# Smartphone pupillometry to predict hematoma expansion in Hemorrhagic stroke patients: A protocol for prospective cohort study

**DOI:** 10.64898/2026.10.13.26365496

**Authors:** Debankur Chakraborty

**Affiliations:** West Bengal University of Health Sciences

## Abstract

**Introduction:** Hematoma expansion is a main cause of early worsening after spontaneous intracerebral hemorrhage (ICH). Many hospitals in low income countries have no invasive pressure monitoring and no automated pupillometer. Staff use the Glasgow Coma Scale (GCS) and a penlight to check the pupils, and pupil changes seen with a penlight usually come late. The speed of pupil constriction may change earlier, and smartphones record video fast enough to measure it.

**Methods and analysis:** This will be a single-centre prospective cohort study of 120 adults with ICH on non-contrast computed tomography (NCCT) who arrive within 24 hours of symptom onset. At admission and every 6 hours until 48 hours, we will record a 6-second smartphone video of each eye reacting to a flash, using an eyecup that blocks ward light. Open-source software will measure the pupil diameter in every frame. The main measure is constriction velocity (CV). The primary outcome is hematoma expansion (an increase of more than 6 mL or more than 33%) on NCCT at 24 hours. Secondary outcomes are a fall of 2 or more GCS points within 48 hours and in-hospital death. The primary analysis estimates the area under the receiver operating characteristic curve for the percentage fall in CV from each patient’s first recording.

**Ethics and dissemination:** Patients or their legal representatives will give written consent. Results will be submitted to a peer-reviewed journal, and the analysis code will be made public.

## Introduction

Spontaneous intracerebral hemorrhage (ICH) causes many stroke deaths and much long-term disability, and most of this burden falls on low and middle-income countries [1]. The hematoma often grows in the first few hours. In an early imaging series, about one in four patients had growth of at least a third within 1 hour of the baseline scan, and 38% had it by 20 hours [2]. Growth is linked to higher mortality and worse functional outcome [3]. It is usually defined as an increase of more than 6 mL or more than 33% [4]. A large trial found that a hemostatic drug reduced growth but did not improve functional outcome [5]. Treatments such as osmotic therapy or surgery depend on noticing early that a patient is getting worse.

Neurocritical care units use invasive intracranial pressure (ICP) monitors and automated infrared pupillometers. The Neurological Pupil index (NPi), calculated by one such device, was introduced as an early sign of rising ICP [6]. A systematic review has summarized the evidence for automated pupillometry in intensive care [7]. Most public hospital wards in low and middle-income countries have neither tool. Staff use the GCS and a penlight, and penlight findings vary between examiners. In 2329 paired assessments, two clinicians agreed only moderately on pupil reactivity (kappa 0.40), size (0.54) and shape (0.62) [8]. In another study, a penlight assessment and an automated pupillometer disagreed on the light reflex in 18% of 406 measurements [9]. A larger pupil on one side, or a fixed pupil, usually appears only after the third cranial nerve is already compressed by herniation.

The pupillomotor parasympathetic fibers run on the outer surface of the oculomotor nerve, and they are thought to be affected by external pressure before the deeper motor fibers. If this is right, pressure from an enlarging hematoma should first slow the constriction of the pupil and lengthen its delay, while the resting pupil size stays close to normal. Automated measures of pupil reactivity have been linked to rising ICP in patients with brain injury [6].

Smartphone cameras record video at 60 to 120 frames per second, which gives one frame every 8 to 17 milliseconds. In healthy people, an iPhone application agreed closely with an infrared camera, with a mean difference of about 6% in the pupil measures [10]. A smartphone flash combined with an infrared camera gave resting pupil diameters comparable to those from a handheld NeurOptics pupillometer [11]. To our knowledge, smartphone pupillometry has not been tested in patients with acute ICH, who are drowsy and need repeated checks on a busy ward.

This protocol describes a study to find out whether serial smartphone recordings of the pupillary light reflex can predict hematoma expansion in patients with ICH on a ward without invasive monitoring.

## Objectives and hypothesis

The primary objective is to estimate how well a fall in pupillary constriction velocity (CV), measured with a smartphone and compared with each patient’s own first recording, predicts hematoma expansion on follow-up NCCT. The secondary objectives are:

1. To describe changes in CV, latency, constriction amplitude and baseline pupil diameter over the first 48 hours, and to compare patients with and without hematoma expansion.
2. To test whether a fall in CV is associated with clinical deterioration (a fall of 2 or more GCS points) within 48 hours after adjusting for the ICH score, and to measure how many hours before deterioration the fall appears.
3. To compare the timing of the smartphone signal with the first abnormal penlight finding recorded by ward staff.

We hypothesize that CV falls and latency lengthens before hematoma expansion or GCS decline becomes apparent, while the resting pupil diameter changes little.

## Methods

### Study design and setting

This is a single-center, prospective, observational cohort study. Patients will be recruited from the acute medicine and neurology step-down wards of a tertiary care hospital. Figure 1 shows the flow of patients and assessments. The study will be reported following the STROBE statement for observational studies [12] and the TRIPOD statement for prediction studies [13].

**Figure 1.**
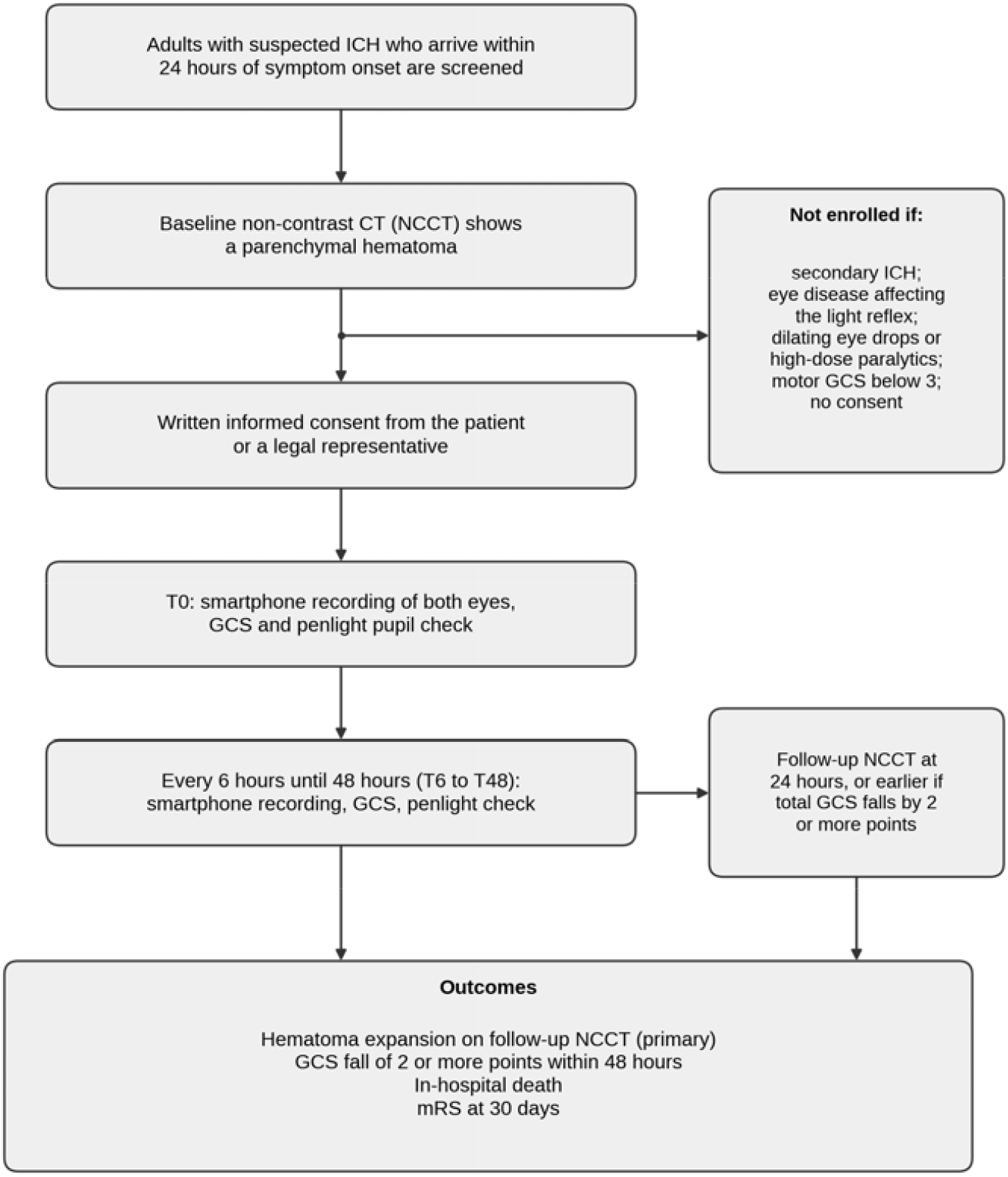
Flow of patients and assessments. ICH, intracerebral hemorrhage; NCCT, non-contrast computed tomography; GCS, Glasgow Coma Scale; mRS, modified Rankin Scale.

### Participants

Consecutive adults with suspected ICH will be screened. Patients are eligible if all of the following apply:

- age 18 years or older;
- arrival within 24 hours of the onset of symptoms;
- a parenchymal hematoma on the baseline NCCT;
- a motor GCS of 3 or more at enrollment;
- written informed consent from the patient or a legally authorized representative. Patients are excluded if any of the following apply:
- secondary ICH from trauma, a vascular malformation (arteriovenous malformation or ruptured aneurysm) or hemorrhagic transformation of an ischemic infarct;
- an eye condition that affects the light reflex, such as past iris trauma, severe bilateral cataract, active anterior uveitis or aniridia;
- topical eye drops that dilate the pupil (for example atropine or tropicamide), or high-dose systemic neuromuscular blocking drugs.

Patients who die or leave the hospital before the first recording will not count toward the 120 participants.

### Smartphone recordings

All recordings will use one smartphone mode. Its rear camera records 1080p video at 60 frames per second, with exposure and focus locked. A 3D-printed eyecup with a silicone rim fits over the camera and the flash. It holds the camera 12 cm from the eye and blocks ward light. The rim rests on the bone around the eye socket and does not touch the eyeball. The eyecup is cleaned with a disinfectant wipe after each patient.

Each recording lasts 6 seconds (Figure 2). The pupil is recorded at rest for 2.0 seconds. The phone LED then switches on for 1.5 seconds. After it switches off, the pupil is recorded for 2.5 seconds while it recovers. Before the flash, the light reaching the eye inside the eyecup should be below 5 lux. The flash should give about 40 lux at the cornea. We will check both values with a lux meter before the first patient and after any change of phone or eyecup. If the LED at its lowest setting is brighter than 40 lux, we will cover it with a diffuser film until the reading is about 40 lux.

**Figure 2.**
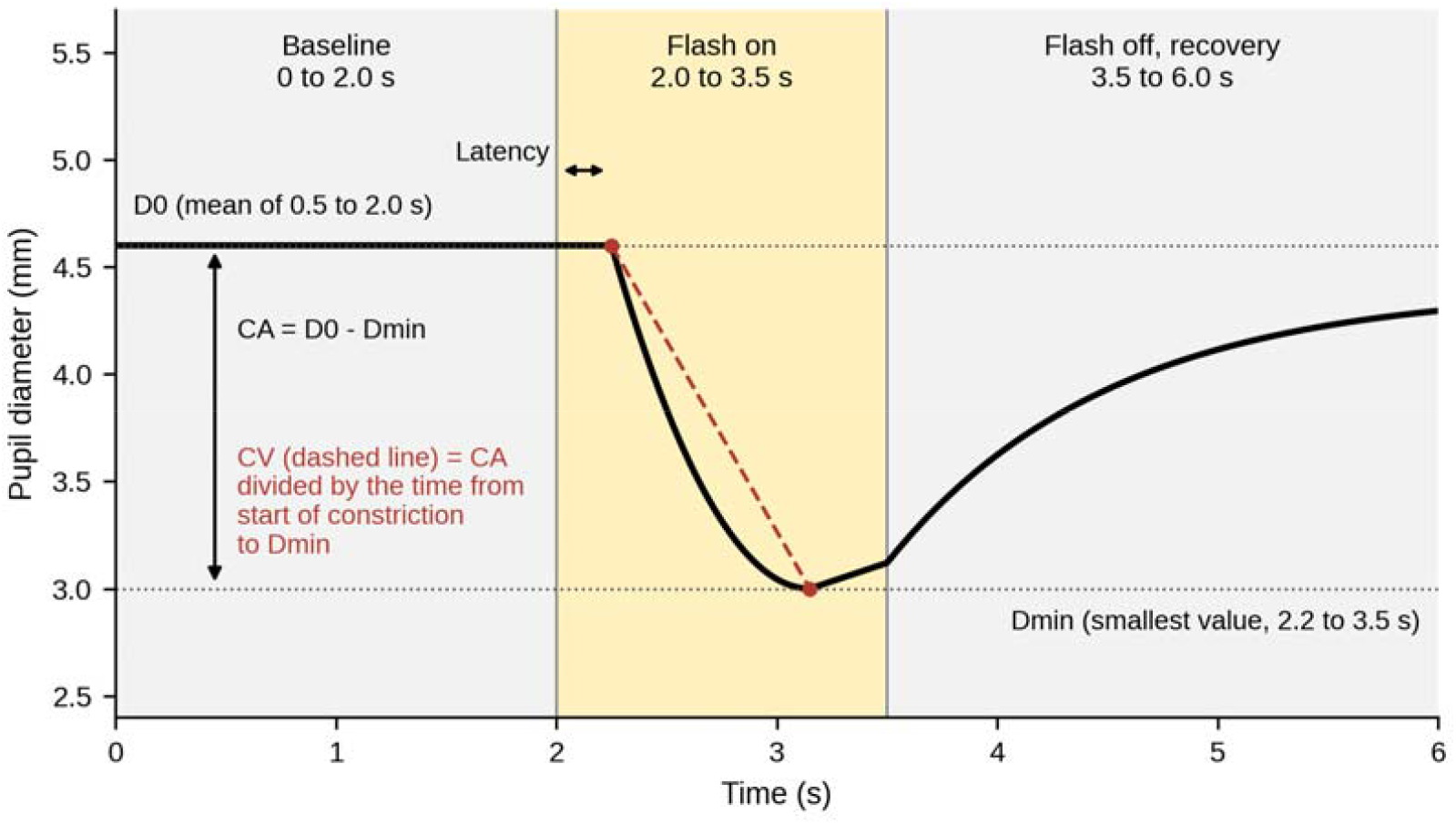
Recording sequence and pupil measures. The curve is a schematic and does not show patient data. D0, baseline diameter; Dmin, minimum diameter; CA, constriction amplitude; CV, constriction velocity.

The patient lies on their back with the head of the bed raised to 30 degrees. If the eye is closed, the researcher holds the upper eyelid open with a thumb on the brow. The right eye is recorded first and then the left eye, with at least 30 seconds between the two so that the pupil can return to its resting size. A failed recording is repeated once. Two trained researchers will make all recordings. Each will complete a 1-hour training session and 10 practice recordings in healthy volunteers before the first patient is enrolled.

The first recording (T0) is made as soon as possible after the baseline NCCT and consent. Recordings are repeated every 6 hours until 48 hours, which gives nine time points (Table 1). Each recording may be made up to 30 minutes before or after its planned time. In the first 20 patients, two recordings of each eye are made 2 minutes apart at T0 to measure repeatability.

**Table 1.** Schedule of assessments. Column headings give the hours after the first recording (T0).

| Assessment | T0 | 6 | 12 | 18 | 24 | 30 | 36 | 42 | 48 | Day 30 |
| --- | --- | --- | --- | --- | --- | --- | --- | --- | --- | --- |
| Written informed consent | X |  |  |  |  |  |  |  |  |  |
| Baseline NCCT (done before T0) | X |  |  |  |  |  |  |  |  |  |
| Smartphone recording, both eyes | X | X | X | X | X | X | X | X | X |  |
| Duplicate recordings (first 20 patients) | X |  |  |  |  |  |  |  |  |  |
| GCS | X | X | X | X | X | X | X | X | X |  |
| Penlight pupil check by ward staff | X | X | X | X | X | X | X | X | X |  |
| Opioids and sedatives in the last 6 hours | X | X | X | X | X | X | X | X | X |  |
| Follow-up NCCT (24 hours after baseline scan, or earlier if GCS falls by 2 or more points) |  |  |  |  | X |  |  |  |  |  |
| mRS (telephone or clinic visit) |  |  |  |  |  |  |  |  |  | X |

### Outcomes

The baseline NCCT is done at presentation. The follow-up NCCT is done 24 hours after the baseline scan, or earlier if the patient deteriorates. Table 2 lists the outcomes.

**Table 2.**
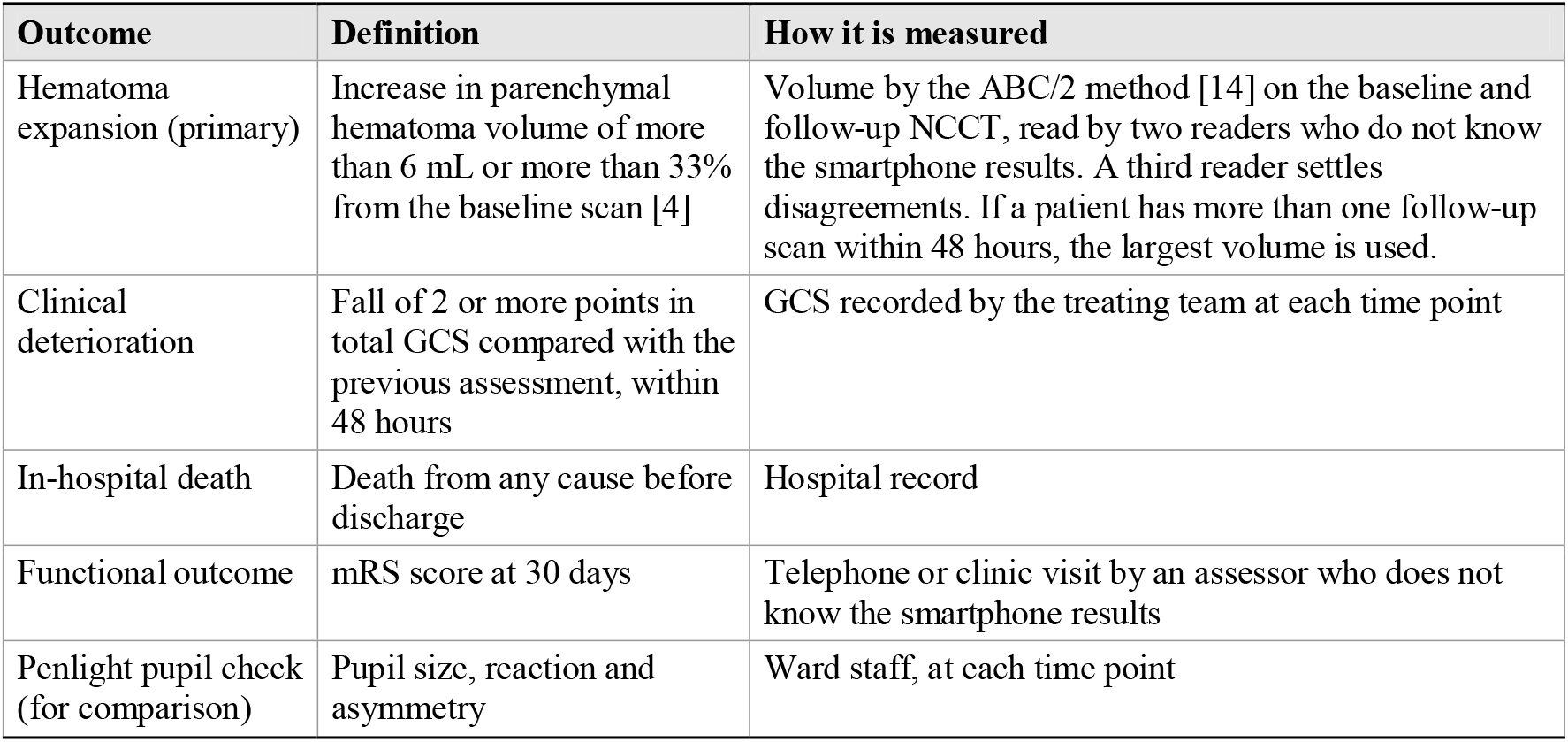
Outcomes, definitions and measurement.

| Outcome | Definition | How it is measured |
| --- | --- | --- |
| Hematoma expansion (primary) | Increase in parenchymal hematoma volume of more than 6 mL or more than 33% from the baseline scan [4] | Volume by the ABC/2 method [14] on the baseline and follow-up NCCT, read by two readers who do not know the smartphone results. A third reader settles disagreements. If a patient has more than one follow-up scan within 48 hours, the largest volume is used. |
| Clinical deterioration | Fall of 2 or more points in total GCS compared with the previous assessment, within 48 hours | GCS recorded by the treating team at each time point |
| In-hospital death | Death from any cause before discharge | Hospital record |
| Functional outcome | mRS score at 30 days | Telephone or clinic visit by an assessor who does not know the smartphone results |
| Penlight pupil check (for comparison) | Pupil size, reaction and asymmetry | Ward staff, at each time point |

### Other data collected

Table 3 lists the other data we will record.

**Table 3.**
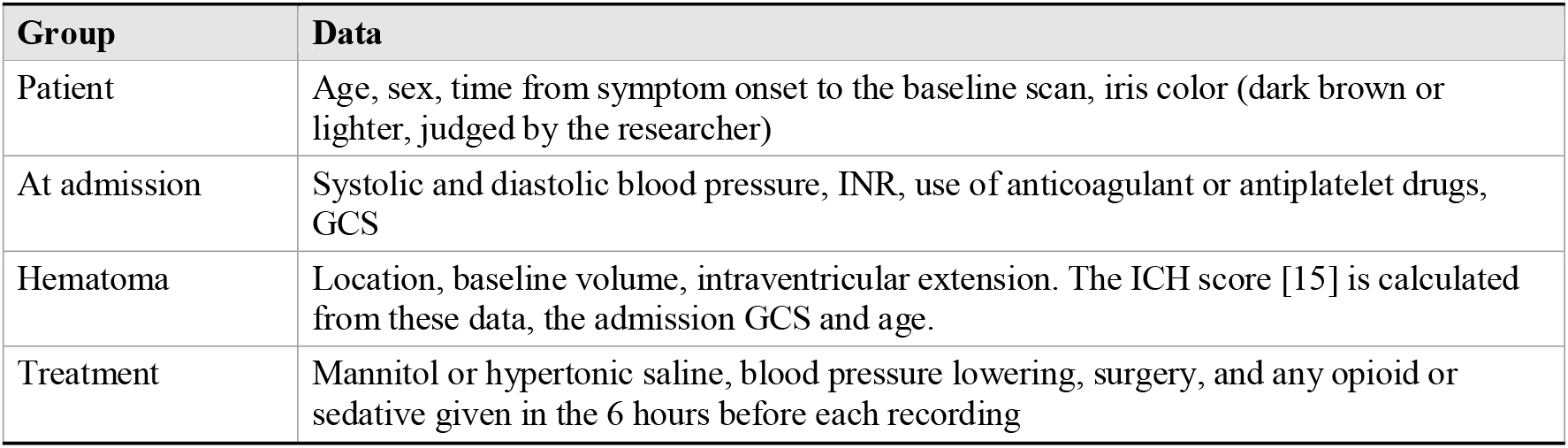
Other data collected.

| Group | Data |
| --- | --- |
| Patient | Age, sex, time from symptom onset to the baseline scan, iris color (dark brown or lighter, judged by the researcher) |
| At admission | Systolic and diastolic blood pressure, INR, use of anticoagulant or antiplatelet drugs, GCS |
| Hematoma | Location, baseline volume, intraventricular extension. The ICH score [15] is calculated from these data, the admission GCS and age. |
| Treatment | Mannitol or hypertonic saline, blood pressure lowering, surgery, and any opioid or sedative given in the 6 hours before each recording |

### Image analysis

The videos will be analyzed with open-source software written in Python, using OpenCV (Figure 3). The main settings are as follows.

1. The first frame in which the mean brightness of the image jumps is taken as the start of the flash. All times are counted from this frame, so software delay does not affect the timing.
2. An open-source facial landmark model finds the eye, and each frame is cropped to the iris.
3. We use the red channel of the color image, because it is expected to give better contrast than grayscale in dark brown irises. Contrast-limited adaptive histogram equalization [17] evens out the brightness, and the reflection of the flash on the cornea is masked. The pupil is separated with an Otsu threshold, and an ellipse is fitted to its edge with the direct least-squares method [18]. Pupil diameter is the mean of the major and minor axes.
4. In the first 20 patients, two readers will outline the pupil by hand in 10 frames per patient (200 frames). We will compare the red channel, the green channel and grayscale with these outlines and keep the one with the smallest mean difference in pupil diameter. This choice is fixed before the main analysis.
5. Pixels are converted to millimeters using the visible horizontal iris diameter, taken as the average for healthy adults, about 11.7 mm [19]. The scale is averaged over the baseline frames of each recording. Percentages do not depend on the scale.
6. A frame is dropped if the aspect ratio of its fitted ellipse differs by more than 20% from the median for that recording, which usually means a blink or a partly closed lid. Gaps of up to 3 frames (50 milliseconds) are filled by linear interpolation. A recording is unusable if more than 20% of the frames between 2.0 and 3.5 seconds are dropped.
7. The diameter curve is smoothed with a 5-frame moving median before any rate of change is calculated.

**Figure 3.**
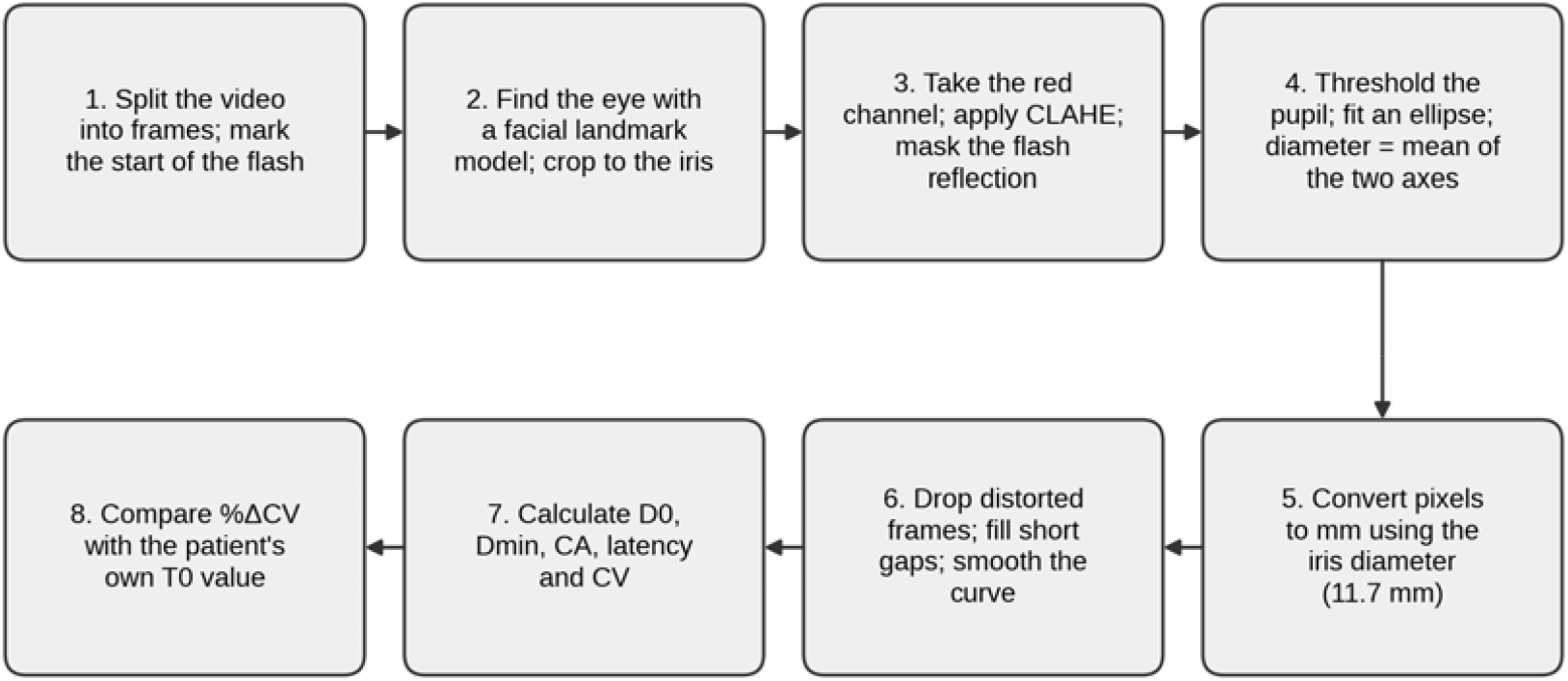
Steps of the image analysis. CLAHE, contrast-limited adaptive histogram equalization; D0, baseline diameter; Dmin, minimum diameter; CA, constriction amplitude; CV, constriction velocity; T0, first recording.

Table 4 defines the measures taken from each usable recording. A researcher who does not know the clinical outcome will check each pupil curve by eye.

**Table 4.** Measures taken from each recording.

| Measure | Definition | Unit |
| --- | --- | --- |
| Baseline diameter (D0) | Mean diameter from 0.5 to 2.0 seconds | mm |
| Minimum diameter (Dmin) | Smallest diameter from 2.2 to 3.5 seconds | mm |
| Constriction amplitude (CA) | $D0 - Dmin$ | mm |
| Percent constriction | $100 \times CA / D0$ | % |
| Latency | Time from the start of the flash to the start of constriction. Constriction starts at the first of 3 consecutive frames in which the smoothed diameter is more than 3 standard deviations below D0 (the standard deviation of the baseline diameter). | s |
| Constriction velocity (CV) | $CA / (\text{time of } Dmin - \text{time of constriction start})$ | mm/s |
| Percentage change in CV (%ΔCV) | $100 \times (CV \text{ at the time point} - CV \text{ at } T0) / CV \text{ at } T0$ | % |

### Sample size

We expect hematoma expansion in about 25% to 38% of patients [2]. If 25% of the 120 patients have expansion, about 30 will have the outcome. If the true area under the curve (AUC) of %^Δ^CV is 0.75, the 95% confidence interval by the Hanley and McNeil method is about 0.64 to 0.86 [20]. We chose 120 patients for this level of precision, and not to give power to a hypothesis test. About 30 events are too few for a prediction model with many predictors [21]. The adjusted analyses therefore include one variable, the ICH score, and are exploratory.

### Statistical analysis

Table 5 lists the planned analysis for each objective.

**Table 5.** Planned analysis for each objective.

| Objective | Outcome | Analysis |
| --- | --- | --- |
| Primary: predict hematoma expansion | Hematoma expansion, yes or no | For each patient, take the largest percentage fall in CV from T0 before the follow-up scan. Estimate the area under the ROC curve with a 95% confidence interval from 2000 bootstrap samples. Choose the cutoff with the Youden index and report sensitivity, specificity and positive and negative predictive values. Also report bootstrap estimates corrected for optimism, because the cutoff is chosen in the same data. |
| Secondary 1: change over time | CV, latency, CA, D0 | Linear mixed-effects model [22] with a random intercept for each patient (and a random slope for time if the data support it). Terms: time, expansion status and their interaction. |
| Secondary 2: clinical deterioration | Time to a fall of 2 or more GCS points | Cox proportional hazards model with $\% \Delta CV$ below the cutoff as a time-varying covariate, adjusted for the ICH score [15]. Lead time is the interval from the first recording below the cutoff to the GCS fall, to the nearest 6 hours. |
| Secondary 3: comparison with penlight | Time of the first abnormal penlight finding | In patients who deteriorate, count how often the first recording below the cutoff came before the first abnormal penlight finding. |
| Secondary 4: death and mRS | In-hospital death; mRS 0 to 2 versus 3 to 6 | Compare the share of patients with $\% \Delta CV$ below the cutoff between those who died and those who survived, and between mRS 0 to 2 and 3 to 6 (Fisher exact test). |
| Secondary 5: feasibility and repeatability | Share of usable recordings; duplicate recordings | Percentage of unusable recordings. Intraclass correlation coefficient and within-patient coefficient of variation of CV from the duplicate recordings. |

Both eyes are recorded. The analysis eye is the one on the same side as the hematoma. For midline or infratentorial hematomas we will use the mean of both eyes. A sensitivity analysis will use the opposite eye. Patients without a follow-up scan, for example because they died early, will be left out of the primary analysis. A sensitivity analysis will count them once as having expansion and once as not having it. The mixed models handle missing recordings under a missing-at-random assumption.

All tests are two-sided with a significance level of 0.05. Secondary analyses are exploratory and will not be adjusted for multiple testing. The analyses will be done in Python.

## Data Availability

All data produced in the present work are contained in the manuscript

## Ethics, consent and data handling

Patients with ICH often cannot give consent, so we will ask a legally authorized representative for written consent and ask the patient again when capacity returns. The eyecup does not press on the eyeball, and each flash lasts 1.5 seconds. Videos show the eye region. They are stored on an encrypted hospital drive under a study number, and only the extracted pupil measurements will leave the research team.

The smartphone results are not given to the treating team, so that they do not change care. There is one exception. If the researcher sees no pupil constriction in one eye, or sees that the resting pupil diameters of the two eyes differ by more than 1 mm when they did not at T0, the researcher tells the treating doctor at once.

### Anticipated problems

Table 6 lists the problems we expect and our response to each.

**Table 6.** Anticipated problems and planned responses.

| **Problem** | **What we will do** |
| --- | --- |
| Restless patients blink and move, which distorts the pupil outline | Drop distorted frames, repeat a failed recording once, and count the unusable recordings as a secondary result. |
| Dark irises give low contrast between pupil and iris in visible light, which is common in South Asian patients | Use the red channel, because longer wavelengths pass through iris pigment more easily, and test it against the green channel and grayscale (Image analysis, step 4). Most phone cameras filter out infrared light, so infrared imaging is not possible with the phone alone. |
| At 60 frames per second, latency can only be measured in steps of 16.7 milliseconds, and normal latency is about 0.2 to 0.3 seconds | Use CV as the main predictor, because it is less affected by the frame interval. Report latency with this limit stated. |
| Phone model and light level can change the stimulus | Use one phone model, check light levels with a lux meter, and do not assume the results apply to other phones. |
| Opioids, sedatives, osmotic therapy and older age change pupil size and reactivity | Record these factors and examine them in sensitivity analyses. |
| One hospital and about 30 events | The result will need external validation and cannot give a cutoff for clinical use. |
| No commercial pupillometer is available for comparison in the same patients | Rely on the repeatability results and on the link with hematoma growth. |

### Dissemination

Results will be submitted to a peer-reviewed journal whatever the findings. The analysis code will be placed in a public repository, and the anonymized pupil measurements will be shared on reasonable request.

## References

1. Krishnamurthi RV, Ikeda T, Feigin VL. Global, regional and country-specific burden of ischaemic stroke, intracerebral haemorrhage and subarachnoid haemorrhage: a systematic analysis of the Global Burden of Disease Study 2017. Neuroepidemiology. 2020;54(2):171–9.

2. Brott T, Broderick J, Kothari R, Barsan M, Sauerbeck L, Zuccarello M, et al. Early hemorrhage growth in patients with intracerebral hemorrhage. Stroke. 1997;28(1):1–5.

3. Davis SM, Broderick J, Hennerici M, Brun NC, Diringer MN, Mayer SA, et al. Hematoma growth is a determinant of mortality and poor outcome after intracerebral hemorrhage. Neurology. 2006;66(8):1175–81.

4. Dowlatshahi D, Demchuk AM, Flaherty ML, Ali M, Lyden PL, Smith EE. Defining hematoma expansion in intracerebral hemorrhage: relationship with patient outcomes. Neurology. 2011;76(14):1238–44.

5. Mayer SA, Brun NC, Begtrup K, Broderick J, Davis S, Diringer MN, et al. Efficacy and safety of recombinant activated factor VII for acute intracerebral hemorrhage. N Engl J Med. 2008;358(20):2127–37.

6. Chen JW, Gombart ZJ, Rogers S, Gardiner SK, Cecil S, Bullock RM. Pupillary reactivity as an early indicator of increased intracranial pressure: the introduction of the Neurological Pupil index. Surg Neurol Int. 2011;2:82.

7. Phillips SS, Mueller CM, Nogueira RG, Khalifa YM. A systematic review assessing the current state of automated pupillometry in the neuroICU. Neurocrit Care. 2019;31(1):142–61.

8. Olson DM, Stutzman S, Saju C, Wilson M, Zhao W, Aiyagari V. Interrater reliability of pupillary assessments. Neurocrit Care. 2016;24(2):251–7.

9. Couret D, Boumaza D, Grisotto C, Triglia T, Pellegrini L, Ocquidant P, et al. Reliability of standard pupillometry practice in neurocritical care: an observational, double-blinded study. Crit Care. 2016;20:99.

10. McAnany JJ, Smith BM, Garland A, Kagen SL. iPhone-based pupillometry: a novel approach for assessing the pupillary light reflex. Optom Vis Sci. 2018;95(10):953–8.

11. Chang LY, Turuwhenua J, Qu TY, Black JM, Acosta ML. Infrared video pupillography coupled with smart phone LED for measurement of pupillary light reflex. Front Integr Neurosci. 2017;11:6.

12. von Elm E, Altman DG, Egger M, Pocock SJ, Gøtzsche PC, Vandenbroucke JP. The Strengthening the Reporting of Observational Studies in Epidemiology (STROBE) statement: guidelines for reporting observational studies. Lancet. 2007;370(9596):1453–7.

13. Collins GS, Reitsma JB, Altman DG, Moons KGM. Transparent reporting of a multivariable prediction model for individual prognosis or diagnosis (TRIPOD): the TRIPOD statement. BMJ. 2015;350:g7594.

14. Kothari RU, Brott T, Broderick JP, Barsan WG, Sauerbeck LR, Zuccarello M, et al. The ABCs of measuring intracerebral hemorrhage volumes. Stroke. 1996;27(8):1304–5.

15. Hemphill JC 3rd, Bonovich DC, Besmertis L, Manley GT, Johnston SC. The ICH score: a simple, reliable grading scale for intracerebral hemorrhage. Stroke. 2001;32(4):891–7.

16. Bradski G. The OpenCV library. Dr Dobb’s Journal of Software Tools. 2000;25(11):120–5.

17. Zuiderveld K. Contrast limited adaptive histogram equalization. In: Heckbert PS, editor. Graphics gems IV. Boston: Academic Press; 1994. p. 474–85.

18. Fitzgibbon A, Pilu M, Fisher RB. Direct least square fitting of ellipses. IEEE Trans Pattern Anal Mach Intell. 1999;21(5):476–80.

19. Rüfer F, Schröder A, Erb C. White-to-white corneal diameter: normal values in healthy humans obtained with the Orbscan II topography system. Cornea. 2005;24(3):259–61.

20. Hanley JA, McNeil BJ. The meaning and use of the area under a receiver operating characteristic (ROC) curve. Radiology. 1982;143(1):29–36.

21. Riley RD, Ensor J, Snell KIE, Harrell FE Jr, Martin GP, Reitsma JB, et al. Calculating the sample size required for developing a clinical prediction model. BMJ. 2020;368:m441.

22. Laird NM, Ware JH. Random-effects models for longitudinal data. Biometrics. 1982;38(4):963–74.

